# Understanding Seizures in Malan Syndrome Through Caregiver Reports: A Cross-Sectional Study

**DOI:** 10.1101/2025.06.13.25329538

**Authors:** Sweta Dubey, Senyene E. Hunter, Christal G. Delagrammatikas, Gloria Pinero, Vimala Elumalai, Muhammad Shahzad Zafar

## Abstract

Malan syndrome is an ultra-rare Overgrowth-Intellectual Disability syndrome caused by pathogenic *NFIX* variants, characterized by intellectual disability, postnatal overgrowth, and dysmorphic features. Seizures in Malan syndrome remain poorly understood. We surveyed caregivers of 53 individuals with Malan syndrome. Overall, 55% had seizures or EEG abnormalities. Seizures occurred in 47%, with 28% experiencing drug-resistant epilepsy. The median seizure onset was at age 3 years. Epilepsy classifications included focal (40%) and unknown-onset tonic-clonic seizures (48%). Generalized tonic-clonic (8%), myoclonic (8%), and epileptic spasms (4%) were also reported. Status epilepticus was common (44%). Valproic acid was the most used anti-seizure medication, with variable efficacy. This study represents the largest cohort to date, providing detailed descriptions of seizures in Malan syndrome, and lays a foundation for future research phenotyping epilepsy in affected individuals. Clinicians should maintain a high suspicion of seizures and monitor closely for status epilepticus in individuals with Malan syndrome.

## INTRODUCTION

Malan syndrome is an ultra-rare overgrowth disorder characterized by macrocephaly, tall stature, intellectual disability, distinct facial features, and behavioral concerns such as aggression and self-injury ^1–4^. Initially classified as Sotos syndrome 2 due to phenotypic similarities, Malan syndrome was recognized as a distinct overgrowth and intellectual disability (OGID) syndrome in 2010, following the identification of heterozygous deletions and point mutations in the nuclear factor I X (NFIX) gene, resulting in *NFIX* haploinsufficiency ^5^. Since its classification, fewer than 100 cases of Malan syndrome have been reported in the literature ^6^, which may reflect under-recognition and under-reporting.

Although intellectual disability and global developmental delay are well-documented in Malan syndrome, other neurological features are less frequently reported. Hypotonia has been described in several studies, and intracranial abnormalities such as hypoplasia of the corpus callosum and optic nerves have been noted ^2^. However, the clinical significance of these findings remains unclear ^1^. While seizures have been mentioned in some broader phenotyping studies of Malan syndrome, detailed information on seizure types, specific electroencephalogram (EEG) findings, or anti-seizure management is largely absent.

Overgrowth–intellectual disability (OGID) syndromes are rare neurodevelopmental disorders characterized by generalized overgrowth (height and/or head circumference ≥2 SD above the mean), intellectual disability, and syndrome-specific clinical features ^7^. Many OGID syndromes, including Malan syndrome, are associated with brain abnormalities, behavioral challenges, and neurodevelopmental conditions such as autism spectrum disorder. Recurrent seizures have been reported in OGID syndromes such as Sotos syndrome, Weaver syndrome, and CHD8-related neurodevelopmental disorder, but their characteristics remain poorly defined ^8^. Recently, the Overgrowth Syndromes Alliance (OSA) emphasized that the burden of seizures on patients and their families is both underrepresented in literature and insufficiently addressed in clinical practice ^8^.

The scarcity of data on seizures in OGID syndromes such as Malan syndrome has led to a limited understanding of seizure types and their management in this population. This study describes seizures in individuals with Malan syndrome, an OGID syndrome, using caregiver-reported data. This study provides a foundation for future research on epilepsy phenotypes in Malan syndrome to improve care for affected children.

## MATERIALS AND METHODS

### Ethical Considerations

The Duke University Health System Institutional Review Board approved this study, and informed consent was obtained from all participants before enrollment.

### Recruitment and Inclusion Criteria

Participants were referred by pediatric neurologists within the Duke University Health System and recruited in collaboration with the Malan Syndrome Foundation. Inclusion criteria included a caregiver-reported genetic diagnosis of Malan syndrome confirmed by genetic testing. Participants without a confirmed genetic diagnosis were excluded from the study. Caregivers provided details of the *NFIX* variants; however, investigators did not review the original genetic reports.

### Data Collection

Data were collected using survey methods similar to those of other published studies assessing epilepsy phenotypes and treatment response ^9^. A validated caregiver-report-based seizure questionnaire was adapted to the Research Electronic Data Capture (REDCap) platform for integration into the Malan syndrome Database ^9,10^. REDCap is a secure, web-based software platform that supports data capture for research studies. The questionnaire captured data on demographics, family history, and medical history, including seizure characteristics (onset, description, duration, and frequency), anti-seizure medications (ASMs) and their perceived effectiveness, other therapies (e.g., ketogenic diet), surgical interventions (e.g., vagal nerve stimulation, resective surgery), EEG and magnetic resonance imaging (MRI) findings, and neurocognitive and psychiatric comorbidities along with their treatments.

Caregiver-reported data was reviewed by epileptologists (MZ and SH). EEG and MRI reports were verified against medical records provided by caregivers when available. Seizure types were classified according to the 2017 International League Against Epilepsy classification system ^11,12^. Caregiver-reported seizure frequency was assessed using a semiquantitative measure in accordance with the validated Epilepsy Learning Health System and Pediatric Epilepsy Learning Health System formats, which evaluate seizure frequency using case report forms ^13,14^. Seizure frequency was categorized and defined as follows: too many to count, at least daily, at least weekly (weekly but not daily), at least monthly (monthly but not weekly), at least yearly (at least once per year but not every month), less than once per year, one single lifetime seizure, and uncertain.

### Data Analysis

Descriptive statistics were used to summarize participant demographics and clinical characteristics. Continuous variables were reported as means and standard deviations for normally distributed data, or medians and interquartile ranges (IQR) for non-normally distributed data. Categorical variables were summarized using frequencies and percentages. Duplicates and incomplete case reports were manually excluded. Not all participants completed every item of the questionnaire. Participants with missing data were excluded from analyses involving the missing variables.

The prevalence of seizures was measured, and the distribution of seizure types, severity, frequency, and treatment patterns was examined. The Kolmogorov-Smirnov test was used to assess the normality of continuous variables. The analysis used the highest reported frequency and duration for participants reporting multiple seizure types. Statistical significance was set at p < 0.05.

## RESULTS

### Clinical Characteristics of Participants (n=53)

Of the 77 participants who consented to the study, 57 completed the standardized questionnaires (Figure 1). After excluding four duplicates, 53 individuals with Malan syndrome were included in the analysis. Participant ages at enrollment and at seizure onset were normally distributed (p = 0.11 and p = 0.12, respectively). The median age of the cohort was 8 years (IQR 5-14 years), which comprised 32% females (n=17) (Table 1). None of the participants reported a family history of Malan syndrome, and only a few (n=4) reported a family history of seizures, limited to second or third-degree relatives.

**Table 1:** Characteristics of Malan Syndrome Patients with and without Seizures. ^1^n (%); Median (IQR) ^2^Participants with variants reported according to American College of Medical Genetics and Genomics guidelines ^3^Deleterious protein changes include frameshifts, nonsense variants, and premature stop codons

| Patient Characteristics |  | All Participants | Participants with Seizures |
| --- | --- | --- | --- |
|  |  | **n=53<sup>1</sup>** | **n=25<sup>1</sup>** |
| Female sex assigned at birth |  | 17 (32%) | 7 (28%) |
| Median Age (years) |  | 8 (IQR 5-14) | 11 (IQR 5.5-14) |
| NFIX variant | Disease-causing variant | 53 (100%) | 25 (100%) |
|  | Deleterious protein changes (n=28)<sup>2,3</sup> | 12 (43%) | 9 (36%) |
|  | *CACNA1A*-containing microdeletions (n=28)<sup>2</sup> | 3 (11%) | 3 (12%) |
| History of Seizures |  | 25 (47%) | 25 (100%) |
| Median Age of Seizure Onset (years) |  | - | 3 (IQR 2-7) |
| Family History | Malan syndrome | 0 (0%) | 0 (0%) |
|  | Seizures | 4 (8%) | 3 (12%) |

**Figure 1.**
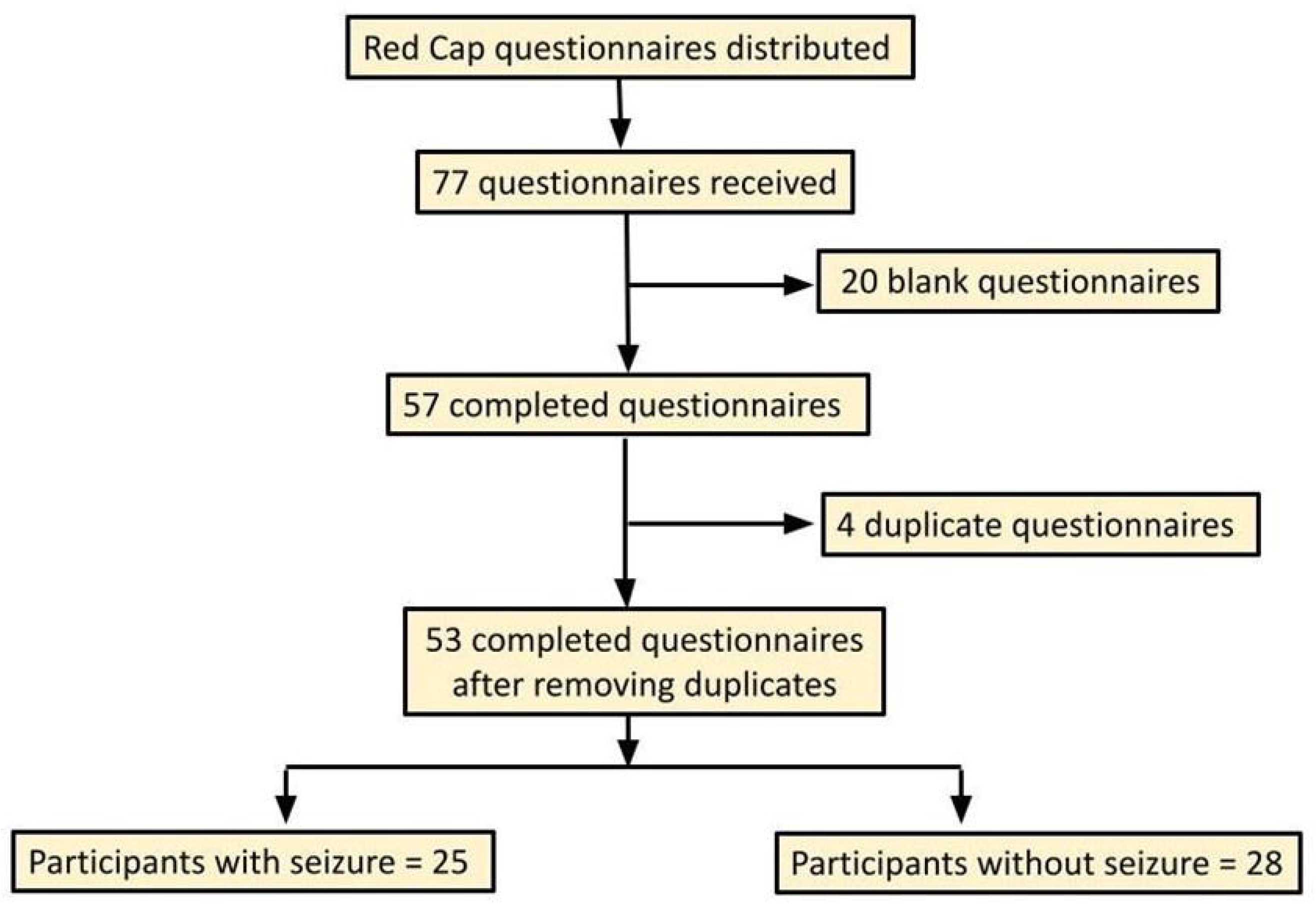
Data Collection and Study Participants. This flow diagram outlines the study’s data collection process and participant inclusion. A total of 77 participants consented to participate, of which 57 completed the standardized questionnaires. Following the removal of four duplicates, 53 patients with a confirmed diagnosis of Malan syndrome were included in the final analysis. The diagram illustrates the steps from initial consent to final inclusion, highlighting the process of data verification.

All participants had genetically confirmed pathogenic or likely pathogenic *NFIX* variants (Table 1). Among these, 64% (n=34) provided specific genetic details, with 53% (n=28) reported according to the American College of Medical Genetics and Genomics (ACMG) guidelines ^15^. Many variants were associated with deleterious protein changes, such as frameshifts, nonsense variants, and premature stop codons (43%, n=12). Pathogenic microdeletions involving the NFIX and CACNA1A genes were reported in 11% (n=3), all of whom had a history of seizures.

### Clinical Characteristics of Participants with Seizures (n=25)

Nearly half (47%, n=25) of the participants had a history of seizures, and 55% (n=29) had either seizures or abnormal EEG findings. Persistent seizures, ongoing at the time of the study, were reported in 28% (n=11) of the cohort. The median age of participants with seizures was 11 years (IQR 5.5-14 years), and the median age of seizure onset was three years (IQR 2-7 years). Three participants reported a family history of seizures, though not in first-degree relatives.

#### Seizure Frequency and Duration

Seizure frequency was classified using the Epilepsy Learning Health System and Pediatric Epilepsy Learning Health System formats, which evaluate seizure frequency using case report forms ^13,14^. Of the 23 participants who provided information on seizure frequency, 39% (n=9) reported seizures occurring at least yearly (Table 2). Seizures occurring at least monthly (22%, n=5), weekly (8.6%, n=2), and daily (17%, n=4) were also noted. Most individuals with seizures (60%, n=15) had persistent seizures at the time of the study, while 12% (n=3) experienced only a single seizure episode in their lifetime. Seizure duration data from 24 participants indicated that most (71%, n=17) had seizures lasting less than five minutes, though 29% (n=7) reported seizures typically lasting five minutes or longer. Notably, 44% (n=11) of those with seizures had at least one episode of status epilepticus.

**Table 2:** Seizure Type, Frequency, and Duration in Individuals with Malan Syndrome and Seizures. ^1^n (%). ^2^Of unknown onset. ^3^Including tonic-clonic, atonic, and myoclonic seizures. ^4^ At least daily = Very frequent seizures: One or more seizures occurring daily. At least weekly = Frequent seizures: One or more seizures occurring over a one-week period. At least monthly = Infrequent seizures: One or more seizures occurring over a one-month period. At least yearly = Rare seizures: One to three seizures over a one-year period.

| **Seizure Types** | **Participants (n=25)<sup>1</sup>** |
| --- | --- |
| Focal onset | 10 (40%) |
| Unknown-onset tonic-clonic | 12 (48%) |
| Generalized tonic-clonic | 2 (8%) |
| Myoclonic<sup>2</sup> | 2 (8%) |
| Multiple types<sup>3</sup> | 5 (20%) |
| Staring spells of unclear etiology | 11 (44%) |
| Infantile/Epileptic Spasm | 1 (4%) |
| Febrile | 6 (24%) |
| **Seizure Frequency<sup>4</sup>** | **Participants (n=23)<sup>1</sup>** |
| At least daily | 4 (17%) |
| At least weekly | 2 (9%) |
| At least monthly | 5 (22%) |
| At least yearly | 9 (39%) |
| Single lifetime episode | 3 (13%) |
| **Typical Seizure Duration** | **Participants (n=24)<sup>1</sup>** |
| < 1 minute | 10 (42%) |
| 1 to 3 minutes | 7 (29%) |
| ≥ 5 Minutes | 7 (29%) |
|  | **Participants (n=25)<sup>1</sup>** |
| History of Status Epilepticus | 11 (44%) |

#### Seizure Types

Seizure types were classified using the 2017 ILAE system based on caregiver responses to a validated epilepsy phenotype survey ^11,12^. The most common seizure types were focal-onset seizures (40%, n=10) and unknown-onset bilateral tonic-clonic seizures (48%, n=12) (Table 2). Generalized-onset tonic-clonic seizures (8%, n=2), myoclonic seizures (8%, n=2), and infantile/epileptic spasms (4%, n=1) were also reported. Multiple seizure types, including tonic-clonic, atonic, and myoclonic seizures, were noted in 20% (n=5) of participants. Febrile seizures occurred in 24% (n=6) of participants, with 20% (n=5) later developing epilepsy. Although 44% (n=11) of individuals reported staring episodes suggestive of absence seizures, some of these participants had a history of focal-onset seizures, and it remains unclear whether absence seizures were formally diagnosed.

### Anti-seizure Management in Participants with Seizures (n=25)

Eighty-four percent (n=21) of individuals with seizures reported using ASMs (Figure 2). The number of ASMs per individual ranged from one to four, with valproic acid being the most commonly used (28%, n=6). Other ASMs included levetiracetam (12%, n=3) and oxcarbazepine (12%, n=3). Combination therapy with two or more ASMs was reported in 24% (n=6) of individuals. Among the 20 participants providing information on ASM efficacy, 67% (n=14) reported well-controlled seizures, while 14% (n=3) had drug-resistant seizures. The ketogenic diet was used by 8% (n=2) of participants and was reported to be effective. One participant with epileptic spasms was treated with corticosteroids, but spasms were refractory, responding only to vigabatrin. Surgical interventions, such as vagus nerve stimulation, were not reported. Although 36% (n=9) of participants reported using supplements such as coenzyme Q10, ubiquinol, levocarnitine, and vitamin B complex, none reported using them specifically for seizure management or neurological concerns.

**Figure 2:**
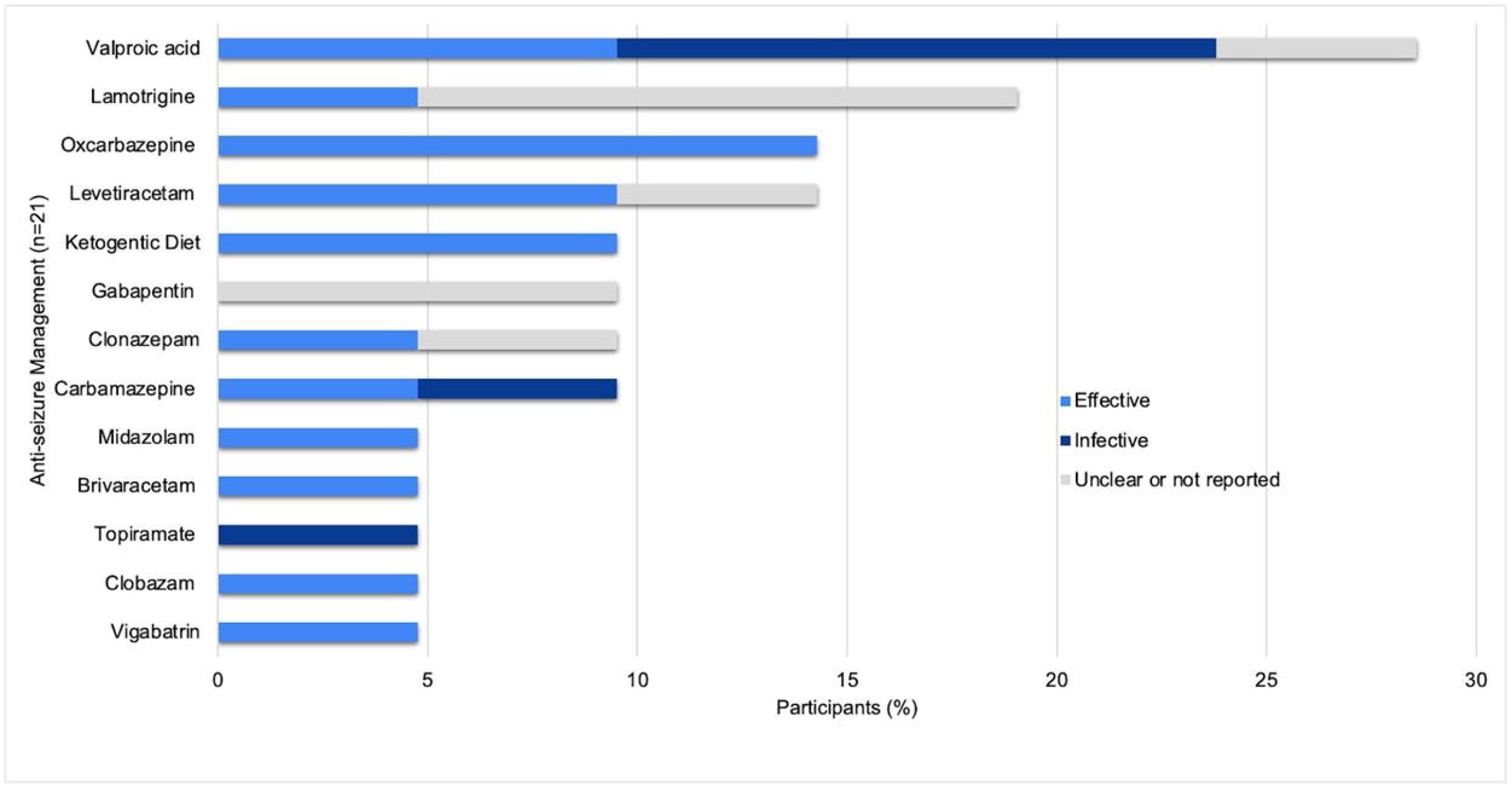
Anti-seizure Management and Perceived Efficacy. The number of participants with seizures who were using anti-seizure medications is presented, along with caregiver-reported perceived efficacy of these treatments. Notably, six participants were on more than one anti-seizure medication, leading to a total number of medication instances that exceeded the number of participants. The colors represent the relative percentages of perceived efficacy: **blue** indicates effective, **dark blue** indicates ineffective, and **gray** indicates unclear or unreported efficacy.

### Diagnostic Testing in Participants with Malan Syndrome (n=53)

#### EEG abnormalities

EEG abnormalities were reported in 61% (n=17) of the 28 individuals who underwent EEG monitoring, with focal epileptiform discharges being the most common abnormality (25%, n=7). These discharges were observed in individuals with focal-onset (n=4) and unknown-onset seizures (n=1), as well as in those without a history of seizures (n=2). Specifically, focal discharges were located in the frontal (4%, n=1), temporal (7%, n=2), and posterior (occipital or parietal-occipital) regions (14%, n=4). Other EEG abnormalities included generalized (4%, n=1) and multifocal discharges (4%, n=1), focal slowing (7%, n=2), spike-and-slow waves (7%, n=2), and rhythmic delta activity (4%, n=1). Diffuse slowing of the background was noted in 32% (n=9), and hypsarrhythmia was observed in one individual with epileptic spasms. Notably, 14% (n=4) of individuals with abnormal EEGs had no history of seizures. Electrographic seizures were captured during EEG monitoring in 7% (n=2) of participants. Ten individuals (36%) had normal EEGs, including three (11%) with a history of seizures.

Among the 25 participants with seizures, 84% (n=21) underwent EEG monitoring. The majority (76%, n=16) had routine EEGs lasting less than one hour, while one participant (4%) had a prolonged EEG lasting up to 12 hours. Continuous inpatient video EEG monitoring was performed on 19% (n=4) of individuals.

#### MRI abnormalities

Brain MRI findings were available for 41 individuals, with abnormalities reported in 61% (n=25). Common findings included prominent ventricles, optic nerve hypoplasia, thinning of the corpus callosum, and nonspecific white matter abnormalities. Specific abnormalities included diffuse abnormal gyration (7%, n=3), a dural defect with herniation of the occipital gyri (2%, n=1), and subependymal heterotopic gray matter (2%, n=1). These specific MRI findings were observed in patients without a history of seizures.

### Neurodevelopment and Neurobehavioral Disorders (n=53) (Table 3)

Developmental delays were common, with speech (81%, n=43), gross and fine motor (79%, n=42), and social delays (38%, n=20) being the most frequently reported. Interventions included speech therapy (77%, n=41), physical or occupational therapy (68%, n=36), and social therapy (7.5%, n=4). Autism spectrum disorder was reported in 13% (n=7), with 6% (n=3) receiving Applied Behavior Analysis therapy. Learning and cognitive delays were prevalent, with 93% (n=40) of respondents indicating that the participant was behind their expected grade level. Additionally, 88% (n=38) had an Individualized Education Plan, and 40% (n=17) were in self-contained classrooms.

**Table 3:** Comorbidities and Developmental Delays and Disabilities in Patients with Malan Syndrome. Footnote: ADHD = attention deficit hyperactivity disorder

| **Comorbid Conditions** | **Number of Participants** |
| --- | --- |
| Anxiety | 29 (55%) |
| Headache | 9 (17%) |
| Sleep disorder | 9 (17%) |
| ADHD | 8 (15%) |
| Depression | 1 (2%) |
| **Developmental Delays and Disabilities** | **Number of Participants** |
| Speech delay | 43 (81%) |
| Gross motor delay | 42 (79%) |
| Fine motor delay | 42 (79%) |
| Social delay | 20 (38%) |
| Autism | 7 (13%) |

Anxiety was reported by 55% (n=29) of participants, with management strategies including fluoxetine, citalopram, risperidone, and sertraline. Other common concerns included attention-deficit/hyperactivity disorder (ADHD) (15%, n=8), significant problems with sleep initiation or maintenance (17%, n=9), and depression (2%, n=1). Headaches were reported in 17% (n=9) and were refractory to treatments such as acetaminophen, verapamil, valproic acid, and in some cases, a ventriculoperitoneal shunt (7.5%, n=4).

## DISCUSSION

This study highlights seizures as a prevalent and clinically important caregiver-reported feature of Malan syndrome. Nearly half of the participants reported seizures, and over half had either seizures or abnormal EEG findings. While prior studies have contributed to the broader understanding of the Malan syndrome phenotype, none have detailed seizure types, frequency, and duration. Findings reported here contribute to the limited literature on seizures as a clinical concern in this rare genetic disorder.

These data align with a recent study involving 16 individuals with Malan syndrome, where 63% had seizures or EEG abnormalities ^1^. In contrast, earlier studies reported lower prevalence estimates. For example, a 2018 study involving 45 individuals found that 18% of participants and 27% of all known patients had seizures or EEG changes ^3^. Similarly, an earlier study reported seizures in approximately 25% of the 20 known patients at that time ^16^. The discrepancy between earlier and more recent findings may reflect increased recognition of seizures in this recently described rare genetic disorder.

Seizure burden varied in this cohort. Some individuals reported a seizure frequency of at least once a year, while others had daily events. Most seizures were brief (less than five minutes), though nearly one-third had prolonged seizures. Notably, nearly half of those with seizures had at least one episode of status epilepticus, suggesting a potentially high risk of status epilepticus in this population.

Focal-onset seizures were most common in this cohort, though myoclonic, generalized tonic-clonic, and epileptic spasms were also reported. Correspondingly, EEG abnormalities varied across the cohort, highlighting the need for more detailed phenotyping of seizure types and associated electrographic features. While prior studies have noted the presence or absence of seizures in Malan syndrome, few, aside from isolated case reports ^17,18,19^, have described seizure characteristics or EEG findings in depth. This large cohort study provides key insights to guide clinical care and future research.

Caregivers reported variability in the ASMs used to treat seizures and in treatment response. While some individuals achieved seizure control, others had drug-resistant epilepsy. Given the limited data on ASM efficacy in this population, further research is needed to identify effective treatment strategies. The high prevalence of seizures and EEG abnormalities in this cohort supports the use of EEG monitoring, as recommended in the proposed minimal dataset for clinical evaluation and management in patients with Malan syndrome ^1^.

Consistent with the previous studies, several comorbid conditions were reported by caregivers, including headaches, ADHD, sleep disorders, anxiety, and developmental delays ^1,2,20^. There is limited literature available regarding characteristics of seizures in OGID to validate our findings ^8^.

This study has several limitations. The reliance on caregiver reports, while valuable, introduces potential biases, including recall bias and subjective interpretation of seizure events. Additionally, the lack of standardized clinical assessments means that some seizure types or events may have been underreported or misclassified. However, this study provides a foundation for future investigations on epilepsy phenotypes in Malan syndrome. Future studies combining caregiver reports with clinical evaluations, EEG data, and genetic analysis may provide a more detailed and accurate characterization of seizures in Malan syndrome. Although the sample size was relatively small, this study represents the largest cohort focusing on seizures in this rare disorder. It is also noteworthy that the majority of the study population was male. Although demographic information on Malan syndrome is limited, the sex distribution in this study does not align with previous reports of sex assigned at birth, which indicates a relatively equal distribution between males and females ^1^.

## CONCLUSION

This study demonstrates that seizures are a prevalent and significant feature of Malan syndrome, an OGID syndrome, contributing to its complex clinical presentation. Enhancing our understanding of the neurological features of Malan syndrome is crucial for improving clinical outcomes for affected individuals and offering better support to their families. Clinicians do not widely recognize Malan syndrome despite its distinctive dysmorphic features. Additionally, the literature on the association between seizures and Malan syndrome has been limited. Clinicians should maintain a high index of suspicion for Malan syndrome when evaluating patients with seizures, particularly those presenting with a consistent clinical phenotype.

## Data Availability

All data produced in the present study are available upon reasonable request to the authors

## ACKNOWLEDGEMENTS

The authors extend their heartfelt thanks to the patients with Malan syndrome and their families for their enthusiastic participation in this study. The authors also gratefully acknowledge the support of the Malan Syndrome Foundation.

## ROLE OF AUTHORS

Study concept and design: Senyene Hunter, Muhammad Zafar

Acquisition, analysis, or interpretation of data: Sweta Dubey, Senyene Hunter, Muhammad Zafar, Christal Delagrammatikas, Vimala Elumalai

Drafting of the manuscript: Sweta Dubey, Senyene Hunter, Muhammad Zafar

Critical revision of the manuscript for important intellectual content: Sweta Dubey, Senyene Hunter, Muhammad Zafar, Christal Delagrammatikas

Obtained study funding: Not Applicable

Administrative, technical, or material support: Gloria Pinero

Study supervision: Muhammad Zafar

## STATEMENTS AND DECLARATIONS

Ethical considerations - The ethics committee approval was obtained from the Duke University Health System Institutional Review Board. The IRB approval/reference number was Pro00105608.

Consent to participate - Written consent was obtained from the caregivers of participating patients.

Consent for publication - Not applicable

Declaration of conflicting interest - All authors declare no conflicting interests.

Funding statement - No funding was obtained for this study.

Data availability - The datasets generated during and/or analyzed during the current study are available from the corresponding author on reasonable request

## Notes

### Competing Interest Statement

The authors have declared no competing interest.

### Funding Statement

This study did not receive any funding

### Author Declarations

Duke University Health System Institutional Review Board provided ethical approval for this work. The IRB approval/reference number was Pro00105608.

## REFERENCES

1. Macchiaiolo M, Panfili FM, Vecchio D, et al. A deep phenotyping experience: up to date in management and diagnosis of Malan syndrome in a single center surveillance report. Orphanet J Rare Dis. 2022;17(1):235.

2. Priolo M. NFIX-Related Malan Syndrome. In: Adam MP, Feldman J, Mirzaa GM, et al., eds. GeneReviews®. University of Washington, Seattle; 1993.

3. Priolo M, Schanze D, Tatton-Brown K, et al. Further delineation of Malan syndrome. Hum Mutat. 2018;39(9):1226–1237.

4. Malan V, Rajan D, Thomas S, et al. Distinct effects of allelic NFIX mutations on nonsense-mediated mRNA decay engender either a Sotos-like or a Marshall-Smith syndrome. Am J Hum Genet. 2010;87(2):189–198.

5. Yoneda Y, Saitsu H, Touyama M, et al. Missense mutations in the DNA-binding/dimerization domain of NFIX cause Sotos-like features. J Hum Genet. 2012;57(3):207–211.

6. Huynh TN, Delagrammatikas CG, Chiriatti L, et al. Natural history in Malan syndrome: survey of 28 adults and literature review. Orphanet J Rare Dis. 2024;19(1):282.

7. Edmondson AC, Kalish JM. Overgrowth Syndromes. J Pediatr Genet. 2015;4(3):136–143.

8. Grens K, Church KM, Diehl E, et al. Epilepsy and overgrowth-intellectual disability syndromes: a patient organization perspective on collaborating to accelerate pathways to treatment. Therapeutic Advances in Rare Disease. 2024;5:26330040241254124.

9. Reutens DC, Howell RA, Gebert KE, Berkovic SF. Validation of a questionnaire for clinical seizure diagnosis. Epilepsia. 1992;33(6):1065-1071.

10. REDCap.

11. Scheffer IE, Berkovic S, Capovilla G, et al. ILAE classification of the epilepsies: Position paper of the ILAE Commission for Classification and Terminology. Epilepsia. 2017;58(4):512–521.

12. Fisher RS, Cross JH, French JA, et al. Operational classification of seizure types by the International League Against Epilepsy: Position Paper of the ILAE Commission for Classification and Terminology. Epilepsia. 2017;58(4):522–530.

13. Donahue MA, Herman ST, Dass D, et al. Establishing a learning healthcare system to improve health outcomes for people with epilepsy. Epilepsy Behav. 2021;117:107805.

14. Grinspan ZM, Patel AD, Shellhaas RA, et al. Design and implementation of electronic health record common data elements for pediatric epilepsy: Foundations for a learning health care system. Epilepsia. 2021;62(1):198–216.

15. Richards S, Aziz N, Bale S, et al. Standards and guidelines for the interpretation of sequence variants: a joint consensus recommendation of the American College of Medical Genetics and Genomics and the Association for Molecular Pathology. Genet Med. 2015;17(5):405–424.

16. Klaassens M, Morrogh D, Rosser EM, et al. Malan syndrome: Sotos-like overgrowth with de novo NFIX sequence variants and deletions in six new patients and a review of the literature. Eur J Hum Genet. 2015;23(5):610–615.

17. Oshima T, Hara H, Takeda N, et al. A novel mutation of NFIX causes Sotos-like syndrome (Malan syndrome) complicated with thoracic aortic aneurysm and dissection. Hum Gen Variation. 2017;4:17022.

18. Bellucco FT, de Mello CB, Meloni VA, Melaragno MI. Malan syndrome in a patient with 19p13.2p13.12 deletion encompassing NFIX and CACNA1A genes: Case report and review of the literature. Mol Genet Genomic Med. 2019;7(12):e997.

19. Kuroda Y, Mizuno Y, Mimaki M, et al. Two patients with 19p13.2 deletion (Malan syndrome) involving NFIX and CACNA1A with overgrowth, developmental delay, and epilepsy. Clin Dysmorphol. 2017;26(4):224–227.

20. Alfieri P, Montanaro FAM, Macchiaiolo M, et al. Behavioral profiling in children and adolescents with Malan syndrome. Front Child Adolesc Psychiatry. 2023;2.

